# A novel decision modeling framework for health policy analyses when outcomes are influenced by social and disease processes

**DOI:** 10.1101/2025.02.21.25322671

**Authors:** Marika M. Cusick, Fernando Alarid-Escudero, Jeremy D. Goldhaber-Fiebert, Sherri Rose

**Affiliations:** Department of Health Policy and Management, Johns Hopkins Bloomberg School of Public Health, Baltimore, Maryland; Department of Health Policy, Stanford School of Medicine, and Stanford Health Policy, Freeman Spogli Institute for International Studies, Standard University, Stanford, California

## Abstract

**Purpose:** Health policy simulation models incorporate disease processes but often ignore social processes that influence health outcomes, potentially leading to suboptimal policy recommendations. To address this gap, we developed a novel decision-analytic modeling framework to integrate social processes.

**Methods:** We evaluated a simplified decision problem using two models: a standard decision-analytic model and a model incorporating our social factors framework. The standard model simulated individuals transitioning through three disease natural history states–healthy, sick, and dead–without accounting for differential health system utilization. Our social factors framework incorporated heterogeneous health insurance coverage, which influenced disease progression and health system utilization. We assessed the impact of a new treatment on a hypothetical cohort of 100,000 healthy, non-Hispanic Black and non-Hispanic white 40-year-old adults. Primary outcomes included life expectancy, cumulative incidence and duration of sickness, and health system utilization throughout a person’s lifetime. Secondary outcomes included costs, quality-adjusted life years, and incremental cost-effectiveness ratios.

**Results:** In the standard model, the new treatment increased life expectancy by 2.7 years for both non-Hispanic Black and non-Hispanic white adults, without affecting racial/ethnic gaps in life expectancy. However, incorporating known racial/ethnic disparities in health insurance coverage with the social factors framework led to smaller life expectancy gains for non-Hispanic Black adults (2.0 years) compared to non-Hispanic white adults (2.2 years), increasing racial/ethnic disparities in life expectancy.

**Limitations:** The availability of social factors data and complexity of causal pathways between factors may pose challenges in applying our social factors framework.

**Conclusions:** Excluding social processes from health policy modeling can result in unrealistic projections and biased policy recommendations. Incorporating the social factors framework enhances simulation models’ effectiveness in evaluating interventions with health equity implications.

**Highlights:**

- Health policy simulation models that ignore social processes may be biased and lead to suboptimal policy recommendations. To address this, we proposed a novel social factors framework to integrate social factors into decision-analytic models for health policy.
- Applying our social factors framework to a simplified example highlighted the potential bias that results from ignoring social factors. In a standard model, a hypothetical new treatment appeared to have no effect on health disparities. However, incorporating our social factors framework demonstrated that this treatment would exacerbate disparities.
- Incorporating a social factors framework into health policy simulation models has particular relevance for evaluating health interventions with equity implications.

## 1 Introduction

Healthcare policymakers face challenging decisions regarding allocating resources to new or existing interventions subject to constrained budgets. Simulation models can offer rigorous evidence on the long-term health and cost implications of these decisions for populations.^1^ Increasingly, policymakers are concerned about how healthcare technologies impact health disparities, which can be examined by assessing the distribution of health outcomes and costs across a population.^2^

Health policy simulation modeling typically focuses on disease natural history to guide policy decisions. However, these models rarely explicitly incorporate the impact of social factors on health outcomes. The field of social epidemiology has developed frameworks to identify and understand the social drivers of health disparities, including patients’ access to healthcare, health-seeking behavior, socioeconomic status, and other structural barriers.^3–5^ Prior research suggests that up to 40% of explainable variation in health outcomes can be attributed to social and economic factors.^6^ Considering and incorporating these factors into health policy simulation models could be critical for informed decision-making.

Existing approaches, such as distributional cost-effectiveness analyses or the use of group-specific model parameters, can estimate the impact of interventions on health disparities. However, these methods alone may be insufficient because they do not include recommendations for how to simulate causal relationships between social factors and health outcomes. We argue that explicitly modeling these relationships in health policy simulation models improves the transparency of model assumptions, strengthens the rigor of quantitative evidence on health equity implications, and enables the evaluation and comparison of both clinical and social interventions.

In this paper, we propose a framework for integrating social factors into health policy decision-analytic models, henceforth referred to as the social factors framework. To demonstrate the value of our framework, we compared model results from a standard model and a model incorporating the social factors framework for a simplified decision problem.

## 2 Methods

### 2.1 Social factors framework

Our social factors framework includes three components: disease natural history, health system utilization, and social factors (Figure 1). In disease natural history, individuals transition through health states specific to a given disease. In the health system utilization component, individuals transition between various levels of healthcare engagement, such as having no regular healthcare access, receiving regular care, being diagnosed, receiving treatment, and discontinuing treatment. An individual’s disease state can impact their health system utilization. For example, individuals in more advanced disease stages may be more likely to be diagnosed or receive treatment. Additionally, health system utilization can impact disease progression. This can occur when, for instance, individuals receiving treatment may experience slowed disease progression or reduced mortality risk. The third modeling component, social factors, includes health insurance status, socioeconomic status, and additional structural barriers that may impact both the natural history of the disease and health system utilization.

**Figure 1:**
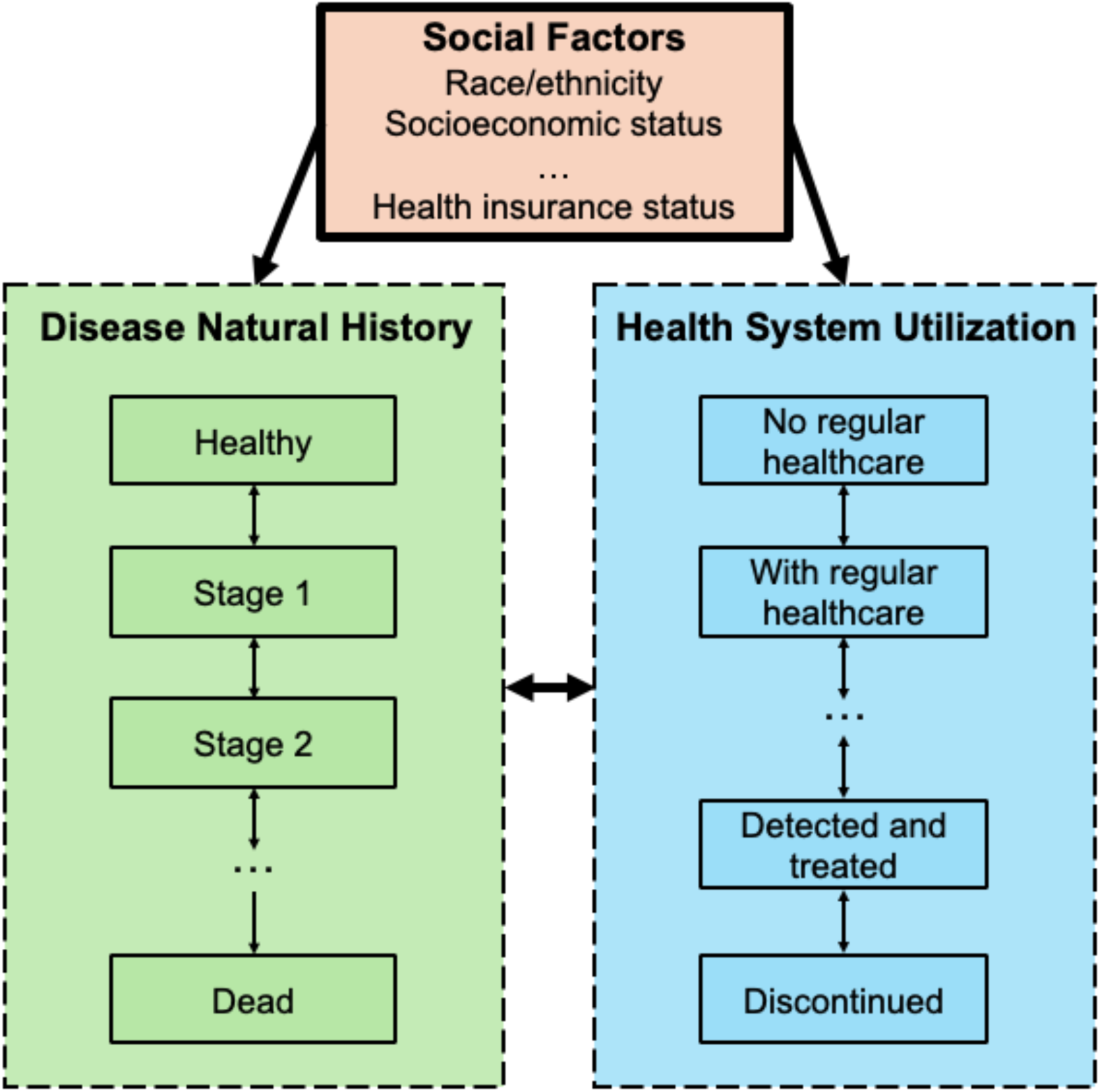
Proposed generalizable social factors framework for including social processes in health policy decision-analytic models.

### 2.2 Simplified decision problem

We compared model results for a simplified decision problem using two state-transition discrete-time decision-analytic microsimulation models: 1) a standard model and 2) a model incorporating our social factors framework (Figure 2). We selected a microsimulation model, an individual-based state-transition model, for analysis as it allows for differentiated simulation pathways according to individual characteristics.^7^

**Figure 2:**
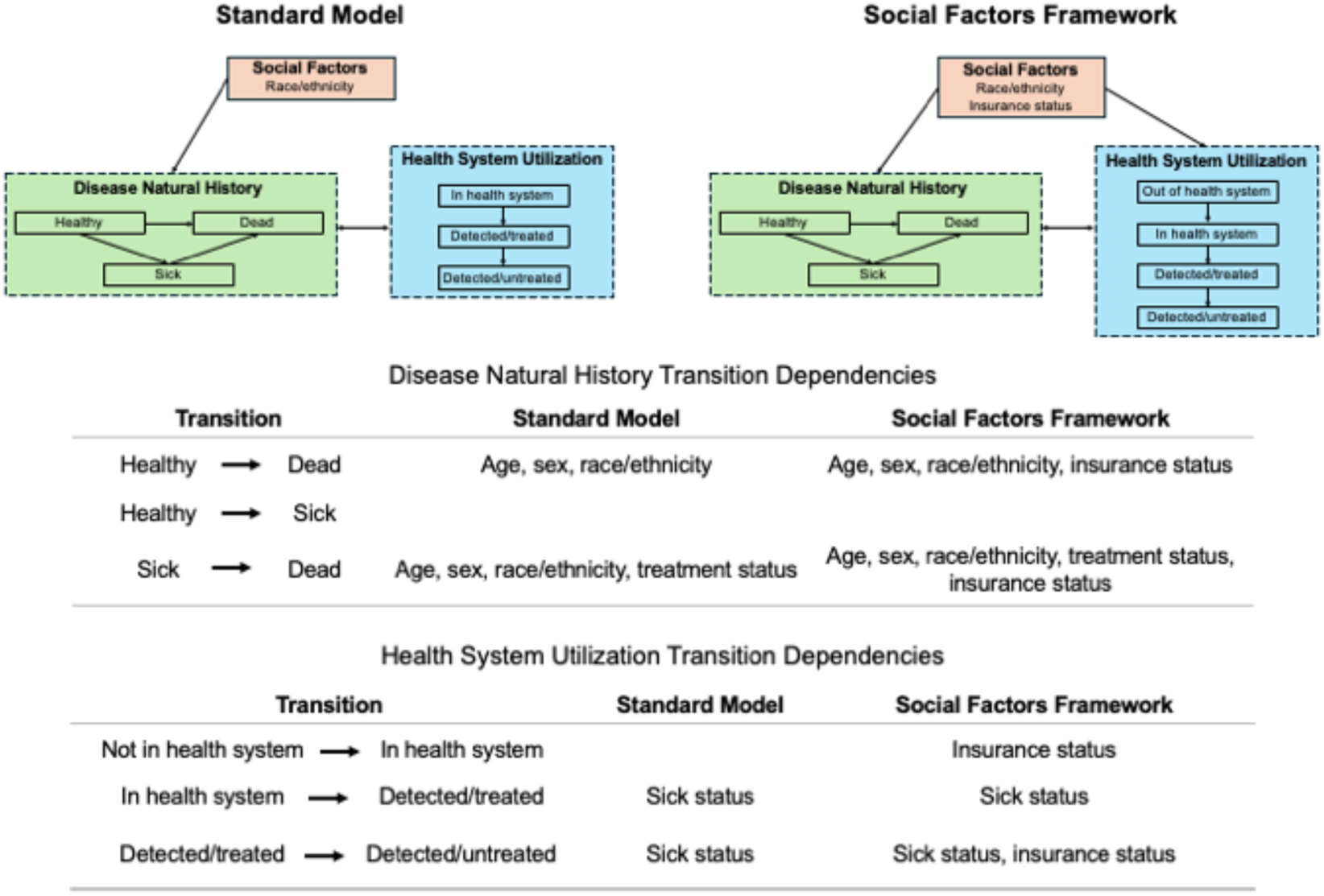
Diagrams and transition dependencies for the standard model and social factors framework for the simplified decision problem^12^

In the standard model, each individual had two characteristics (age and race/ethnicity). As individuals aged, they transitioned through three disease natural history states: healthy, sick, and dead. The standard model incorporated racial and ethnic differences in mortality using age-sex-race-ethnicity-specific U.S. life table rates.^8^ We assumed all individuals were in the health system with regular healthcare access and not detected or treated for the disease. Sick individuals could be detected and treated for the disease and subsequently discontinue treatment.

To incorporate the social factors framework, we modified the standard model by adding several new components. First, we added a social factor, health insurance status. Based on evidence from prior studies, we assumed health insurance status impacted baseline mortality rates and access to regular healthcare.^9–12^ We adjusted age-sex-race-ethnicity-specific U.S. life table mortality rates by insurance status assuming a constant race/ethnicity-specific prevalence of uninsured individuals and a hazard ratio for increased mortality risk among those uninsured.^9,10^

Insurance status also impacted access to routine healthcare and adherence to treatment. In the model with the social factors framework, we added a health system utilization state, “not in health system,” which represented individuals without access to routine healthcare. From the outset, uninsured individuals were less likely to be in the health system with routine access to healthcare compared to those with health insurance coverage.^11,12^ Details for estimating these probabilities are provided in Section 2.3. Individuals not in the health system may enter the health system and gain regular healthcare access over time. Additionally, we assumed that uninsured sick individuals discontinued treatment at faster rates compared to those with insurance. Figure 2 provides a visual comparison of the two models and the transition probability dependencies.

### 2.3 Population and intervention

We simulated a cohort of 100,000 healthy, non-Hispanic Black and non-Hispanic white 40-year-old men and women. We estimated the sex and racial/ethnic group distribution of the simulated cohort, representative of the U.S. population, using a cohort of survey participants from the U.S. National Health and Nutrition Examination Survey (NHANES) 2013-2018 data.^13^ The survey participants were aged between 35 and 45 years, self-identified as either non-Hispanic Black or non-Hispanic white, and self-identified as either female or male.

For our social factors framework, we estimated additional quantities: the proportions of adults with health insurance coverage by race/ethnicity and the proportions of adults with routine healthcare access by insurance status. Insurance coverage proportions by race/ethnicity were estimated with the fraction of individuals in each racial/ethnic group who responded “yes” to the NHANES survey question “Are you covered by health insurance or some other kind of healthcare plan?” Given the stylized nature of the model, for simplicity, we assumed insurance status remained constant over the lifetime. Routine healthcare access proportions were estimated using the fraction of individuals with and without health insurance coverage who responded “yes” or “there is more than one place” to the NHANES survey question, “Is there a place that you usually go when you are sick or you need advice about your health?”

Our simplified decision problem involved evaluating the impact of adding a hypothetical new treatment to standard of care. Without treatment, the sickness increased an individual’s mortality rate by a factor of 4 (i.e., a hazard ratio of 4). An existing standard of care treatment halved the mortality rate from sickness. The new treatment eliminated disease-specific excess mortality such that patients progressed to death at their baseline mortality rates. Base case model parameters are provided in Table 1. We assumed all parameters were fixed and did not introduce parameter uncertainty.

**Table 1:** Base case model parameters for standard model and social factors framework ^3^.

| Parameter |  | Standard Model | Social Factors Framework |
| --- | --- | --- | --- |
| *Disease natural history annual transition probabilities* |  |  |  |
| Healthy | Dead | Age-sex-race-ethnicity-specific U.S. life tables<sup>8</sup> | Age-sex-race-ethnicity-insurance-specific U.S. life tables<sup>8</sup> |
| Healthy | Sick | 0.05 | 0.05 |
| *Increased mortality risk* |  |  |  |
| Hazard ratio of death (sick vs. healthy) |  | 4.0 | 4.0 |
| *Health system transition annual probabilities* |  |  |  |
| Out of health system | In health system | 0.05 | 0.05 |
| In health system | Detected/treated | 0.20 | 0.20 |
| Detected/treated | Detected/untreated | 0.02 | 0.02 |
| *Hazard ratios on risk of health system transitions (uninsured vs. insured)* |  |  |  |
| Out of system | In system |  | 0.20 |
| In system | Detected/treated |  | 0.20 |
| Detected/treated | Detected/untreated |  | 5.0 |
| *Treatment effectiveness hazard ratios on mortality* |  |  |  |
| Standard of care |  | 0.50 | 0.50 |
| Standard of care and new treatment |  | 0.25 | 0.25 |
| *Mortality adjustments by insurance status* |  |  |  |
| Hazard ratio for increased mortality among uninsured adults |  |  | 1.4<sup>9</sup> |
| Prevalence of uninsured across nonelderly | Non-Hispanic Black |  | 10%<sup>10</sup> |

<sup>3</sup> Parameters without citations are assumed parameters.
<sup>4</sup> We estimated the proportion of our cohort in the health system state using the National Health and Nutrition Examination Survey question for routine place to go for healthcare.

|  |  |  |  |
| --- | --- | --- | --- |
| population (0-64y) |  |  |  |
|  | Non-Hispanic white |  | 7%<sup>10</sup> |
| *Cohort characteristics*<sup>13</sup> |  |  |  |
| Sex | Female | 52%<sup>13</sup> | 52%<sup>13</sup> |
|  | Male | 48%<sup>13</sup> | 48%<sup>13</sup> |
| Race/ethnicity | Non-Hispanic Black | 18%<sup>13</sup> | 18%<sup>13</sup> |
|  | Non-Hispanic white | 82%<sup>13</sup> | 82%<sup>13</sup> |
| Insurance coverage | Non-Hispanic Black |  | 76%<sup>13</sup> |
|  | Non-Hispanic white |  | 87%<sup>13</sup> |
| In health system<sup>4</sup> | Adults with insurance |  | 86%<sup>13</sup> |
|  | Adults without insurance |  | 57%<sup>13</sup> |
| *Cost-effectiveness analysis parameters* |  |  |  |
| State-specific quality-of-life weights | Healthy | 1.0 | 1.0 |
|  | Sick | 0.7 | 0.7 |
| Annual state-specific costs | Healthy | \$100 | \$100 |
| | Sick | \$500 | \$500 |
| Annual treatment costs | Standard of care | \$200 | \$200 |
| | New treatment | \$6,000 | \$6,000 |

### 2.4 Outcomes and analysis

Our main analysis focused on health outcomes including life expectancy, cumulative incidence of sickness, years spent in the sick state, cumulative incidence of being diagnosed and treated, and years spent on treatment over the lifetime horizon. For all outcomes, we reported mean estimates and 95% confidence intervals based on the Monte Carlo standard errors (MCSEs) across the standard model and the model incorporating our social factors framework.

As a secondary analysis, we conducted an illustrative cost-effectiveness analysis using both models from the healthcare sector perspective in accordance with the Second Panel on Cost-Effectiveness in Health and Medicine.^2^ Table 1 shows parameters for the illustrative cost-effectiveness analysis. State-specific quality-of-life weights and costs differed by disease natural history states. Treatment costs for the standard of care and the new treatment accrued annually for individuals while they were on treatment. We computed mean and MCSE estimates of discounted quality-adjusted life years (QALYs), costs, and incremental cost-effectiveness ratios (ICERs), all discounted at 3% annually.

### 2.5 Sensitivity analysis

We conducted two sensitivity analyses for the simplified decision problem. First, we compared results from the standard model and the model incorporating the social factors framework for an alternative sickness characterized by lower disease prevalence, increased mortality risk, higher costs, and lower associated quality of life. The modified parameters for this scenario are specified in the Supplement (Table S1).

Second, using base case model sickness parameters (Table 1), we increased the costs associated with sickness and the new treatment for individuals without health insurance (Table S2). This reflects evidence that uninsured individuals have higher out-of-pocket payments.

## 3 Results

Out of the 100,000 simulated individuals, 18% were non-Hispanic Black. In the model incorporating our social factors framework, non-Hispanic Black adults had lower rates of insurance coverage (77%) compared to non-Hispanic white adults (87%). Consequently, a smaller proportion of non-Hispanic Black adults started in the health system (80%) compared to non-Hispanic white adults (82%).

### 3.1 Health outcomes

#### 3.1.1 Life expectancy

Under the standard model, non-Hispanic Black 40-year-old adults receiving the standard of care lived 28.9 years (95% CI: 28.7, 29.1) (Table 2). In comparison, non-Hispanic white 40-year-old adults lived, on average, 3.9 years (95% CI: 3.7, 4.1) longer than non-Hispanic Black adults. Adding the new treatment increased average life expectancy for both non-Hispanic Black and non-Hispanic white adults by 2.7 years, resulting in no changes to the racial/ethnic disparity in life expectancy.

**Table 2:**
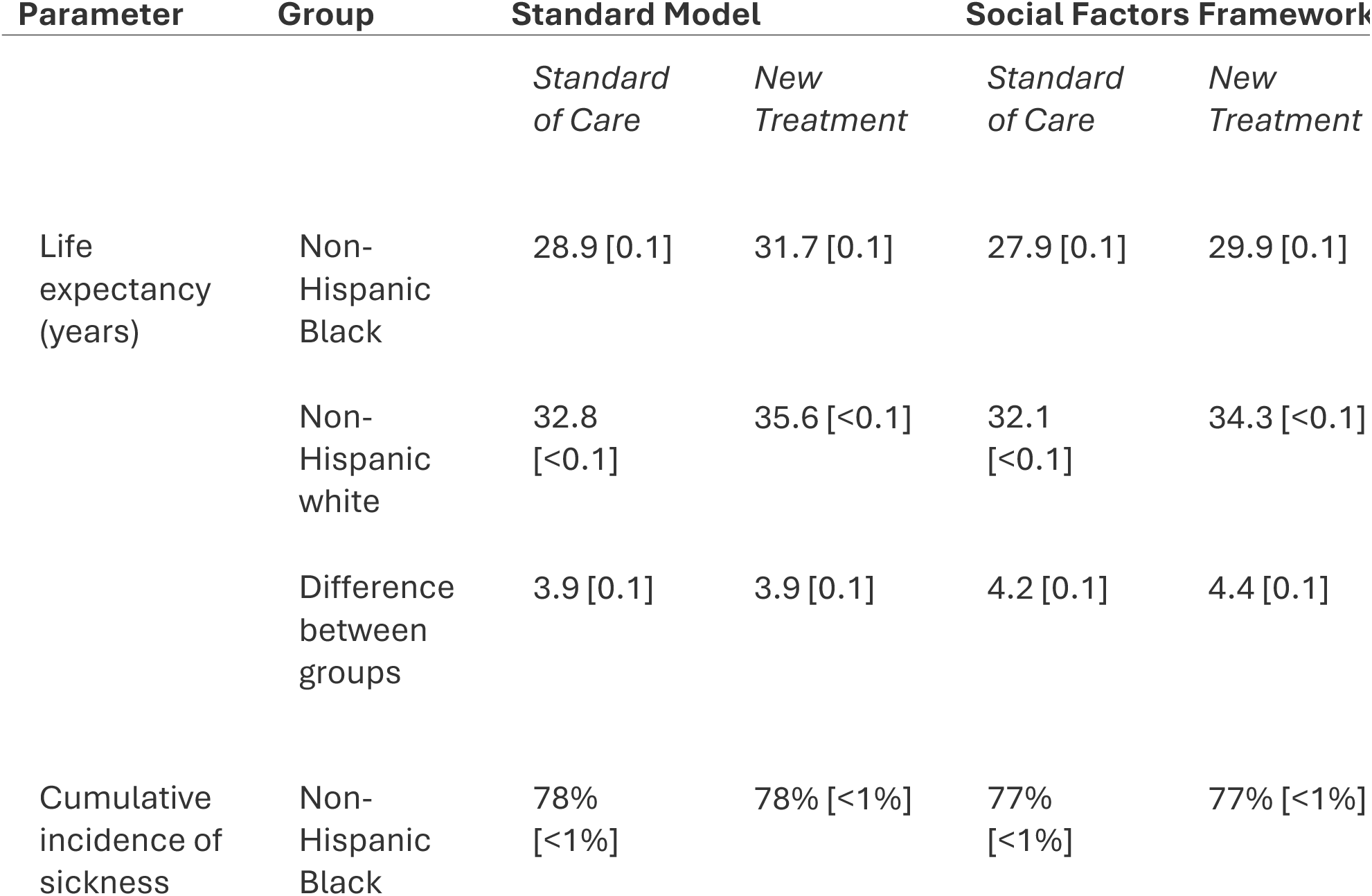
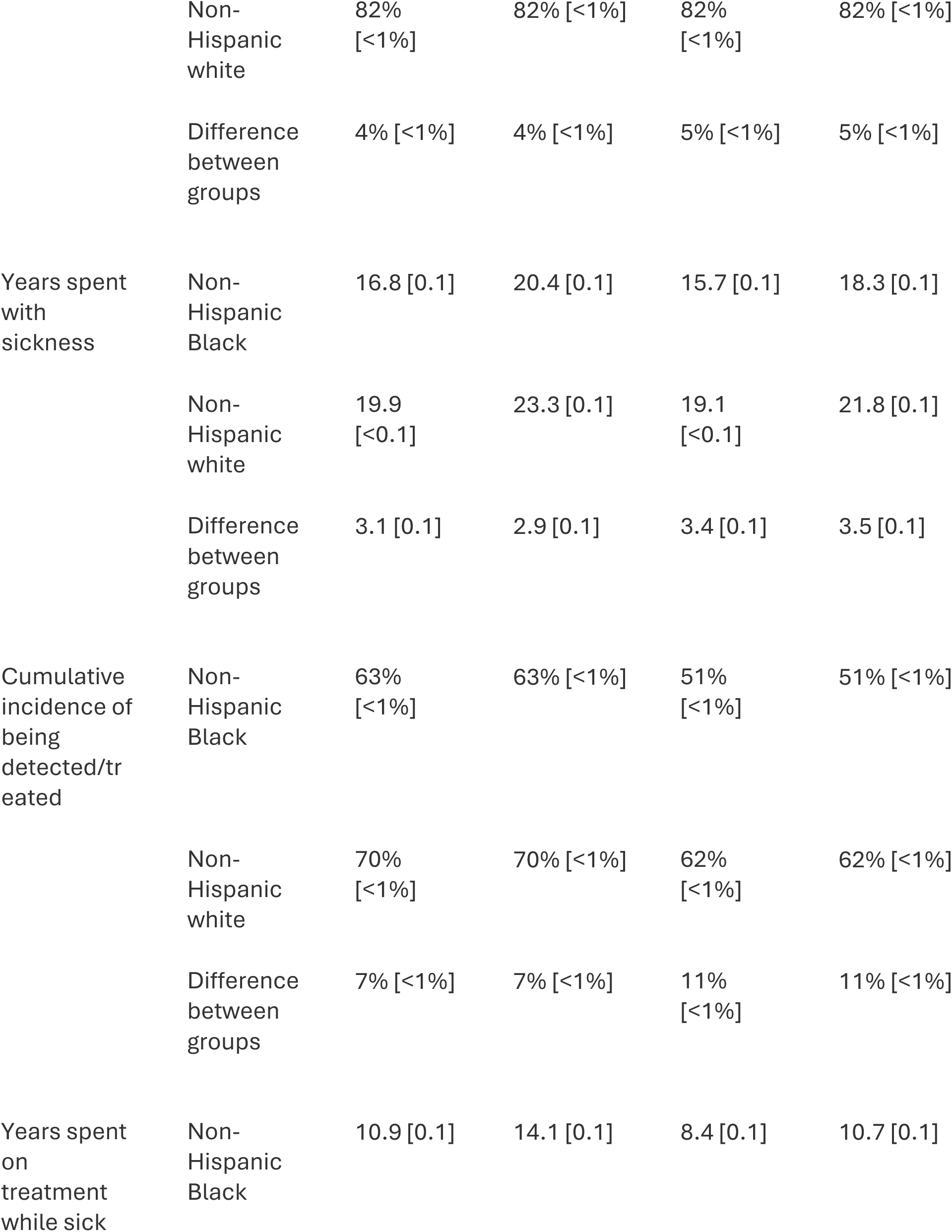
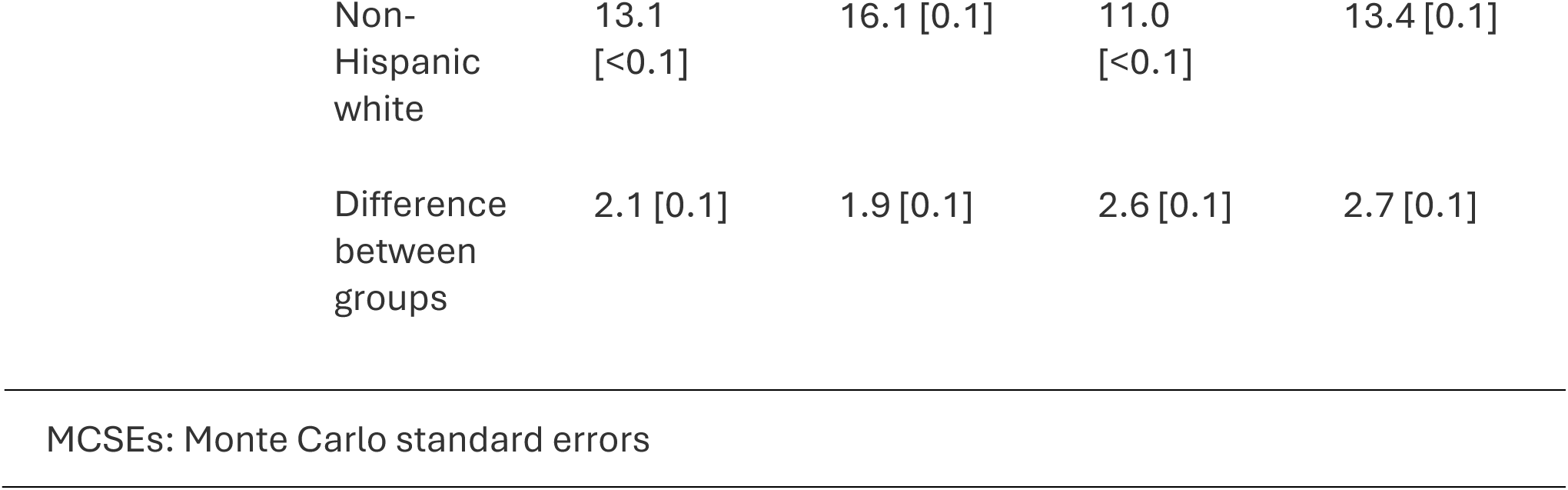
Health outcomes results for standard model and social factors framework (Means [MCSEs])

Using the social factors framework, which accounts for differences in baseline health insurance coverage, the racial/ethnic disparity in life expectancy under the standard of care was 0.3 years greater than in the standard model. Life expectancy gains from the new treatment were also smaller. This effect was more pronounced for non-Hispanic Black adults who gained 2.0 years (95% CI: 1.9, 2.1), compared with 2.2 years (95% CI: 2.2, 2.3) for non-Hispanic white adults. With the social factors framework, adding the new treatment to the standard of care widened the racial/ethnic disparity in life expectancy between non-Hispanic Black and non-Hispanic white adults by 0.2 years.

#### 3.1.2 Cumulative incidence of sickness

The lifetime cumulative incidence of sickness was 78% (95% CI: 77%, 78%) and 82% (95% CI: 82%, 83%) for non-Hispanic Black and non-Hispanic white adults, respectively, with the standard of care in the standard model. Because individuals with sickness cannot be cured, gains in the number of years spent sick were years of successful management and survival of the disease. With the new treatment added to standard of care, non-Hispanic Black adults spent an additional 3.5 years sick (95% CI: 3.4, 3.6), compared with 3.3 years (95% CI: 3.3, 3.4) for non-Hispanic white adults. The racial/ethnic difference in years spent sick narrowed between non-Hispanic Black adults and non-Hispanic white adults by 0.2 years.

In the social factors framework, the lifetime cumulative incidence of sickness was 77% (95% CI: 77%, 78%) and 82% (95% CI: 82%, 82%) for non-Hispanic Black and non-Hispanic white adults, respectively. Compared to the standard model, the average gain in the number of years spent sick was lower for both groups: non-Hispanic Black adults gained 0.9 fewer years sick, and non-Hispanic white adults gained 0.6 fewer years sick. The racial/ethnic difference in the average number of years spent sick increased by 0.1 years with the new treatment.

### 3.2 Health system utilization

All individuals started in the health system with regular healthcare and were eligible for detection and treatment for sickness in the standard model. The cumulative lifetime incidence of being detected and treated when receiving the new treatment was 69% (95% CI: 69%, 69%) across the cohort, with 63% (95% CI: 62%, 64%) and 70% (95% CI: 70%, 71%) for non-Hispanic Black and non-Hispanic white adults, respectively. When receiving the new treatment, non-Hispanic Black adults and non-Hispanic white adults both spent on average 69% of their sick years on the new treatment.

For the social factors framework, 18% of individuals started outside of the health system. Therefore, the cumulative lifetime incidence of being detected and treated was lower than that of the standard model by 9% at 60% (95% CI: 59%, 60%) (Figure 3 (a)). The incidence of being detected and treated was 12% and 9% lower compared to the standard model for non-Hispanic Black and non-Hispanic white adults, respectively. These differences were driven by differences in health insurance coverage, which influenced transitions into the health system and access to treatment (Figure S1). Individuals with insurance had a higher cumulative incidence of being detected and treated at 66% (95% CI: 66%, 67%) throughout their lifetime, thus benefiting more from the new treatment, compared to individuals without insurance at 22% (95% CI: 21%, 22%) (Figure 3 (b)).

**Figure 3:**
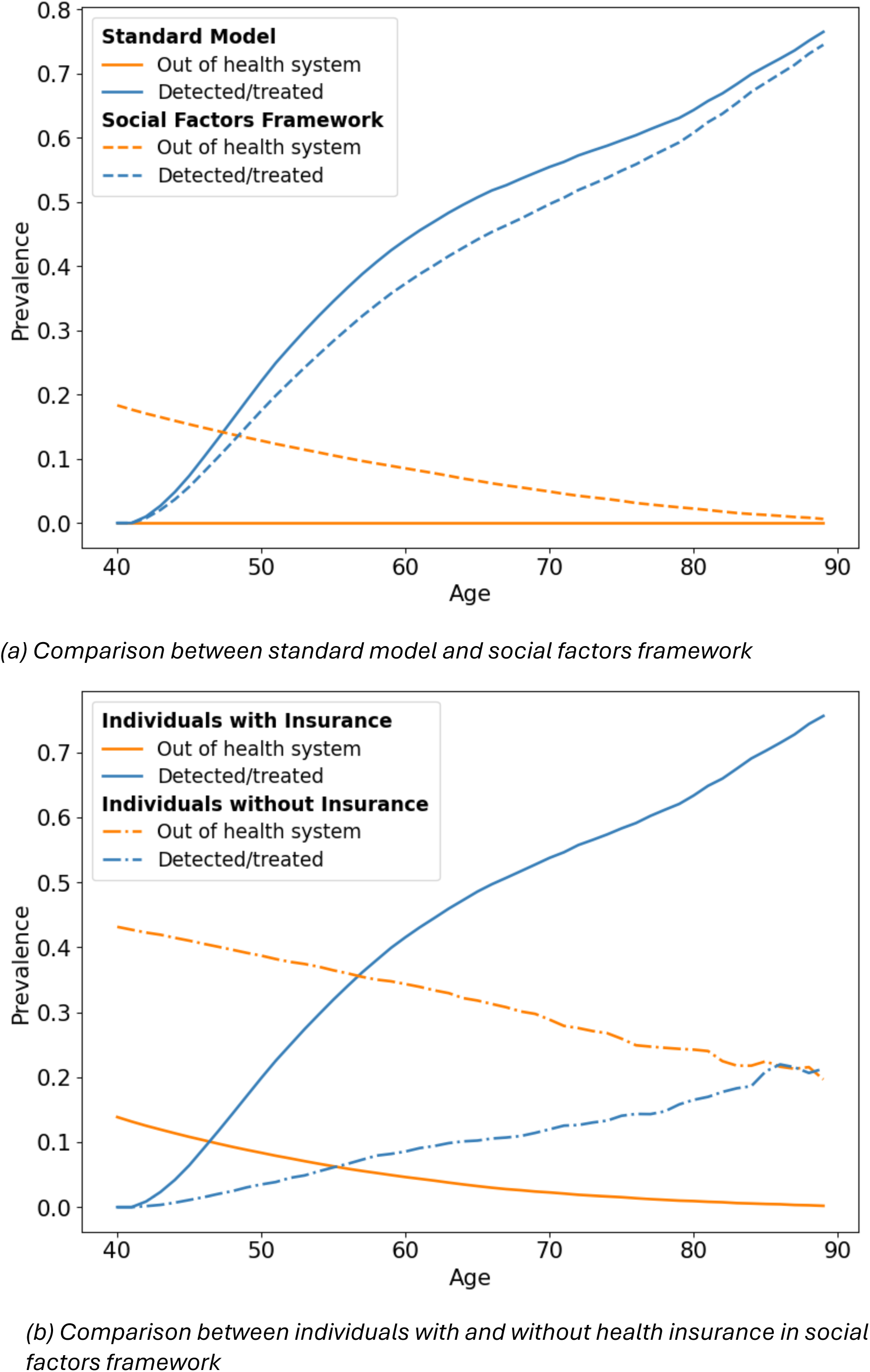
Model trace of simulated cohort across health system utilization states over 60 years.

Additionally, non-Hispanic Black adults and non-Hispanic white adults spent a lower percentage of their sick years on the new treatment in the social factors framework implementation compared to the standard model. Non-Hispanic Black and non-Hispanic white adults spent 59% and 61% of their years sick on treatment, respectively. After accounting for heterogeneity in health insurance coverage with the social factors framework, non-Hispanic white patients spent a greater proportion of sick years on the new treatment (3 percentage points) compared to non-Hispanic Black adults.

### 3.3 Cost-effectiveness analysis

Herein, we summarize differences in our cost-effectiveness analyses. Discounted expected lifetime healthcare costs when receiving the standard of care in the standard model were $5,800 (95% CI: $5,800, $5,900) for a 40-year-old non-Hispanic Black adult (Table S3). Adding the new treatment, which costs $6,000 per year, increased lifetime healthcare costs by $31,400 and yielded an average gain of 0.7 QALYs compared to standard of care. For non-Hispanic white adults, adding the new treatment led to a larger cost increase ($6,800) and smaller QALYs gains (0.6) compared to non-Hispanic Black adults. As a result, the ICER for adding the new treatment was higher for non-Hispanic white adults ($57,100 per QALY gained) compared to non-Hispanic Black adults ($46,200 per QALY gained) (Table S4, Table S5).

The new treatment resulted in lower QALY gains and healthcare cost increases in the social factors framework due to reduced access to treatment, as observed in Section 3.2. These changes were more pronounced for non-Hispanic Black adults, who had lower health insurance coverage compared to non-Hispanic white adults (Figure 4). For non-Hispanic Black adults, the lifetime healthcare costs and QALYs increased by $23,600 and 0.5, which were $7,700 and 0.2 lower than when computed in the standard model. For non-Hispanic white adults, the new treatment resulted in $6,200 less in additional healthcare costs and 0.1 fewer QALYs compared with the standard model.

**Figure 4:**
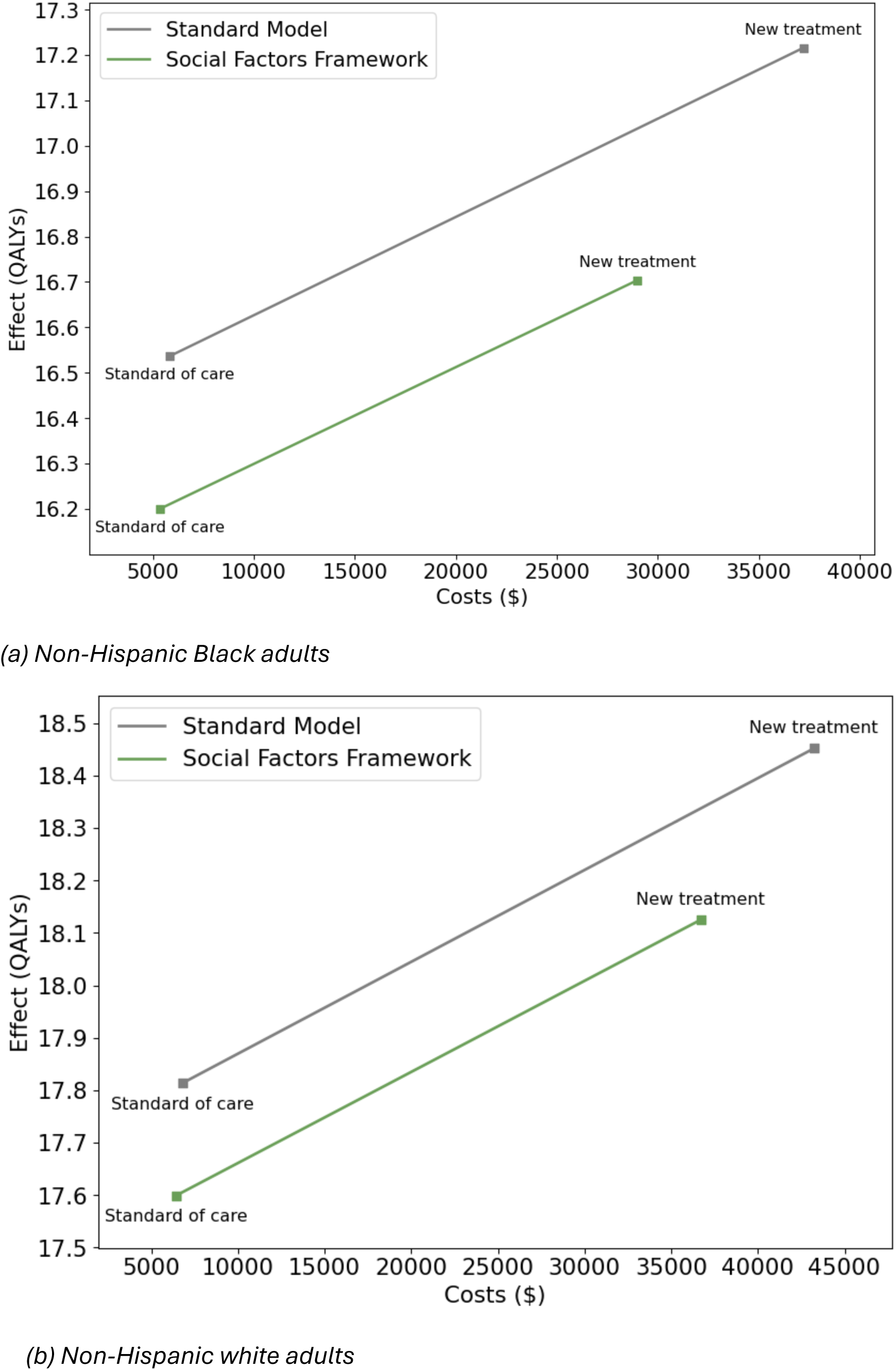
Cost-effectiveness planes.

The ICERs for adding the new treatment were less favorable when using the social factors framework compared to the standard model. Adding the new treatment cost $46,900 and $57,600 per QALY gained for non-Hispanic Black and non-Hispanic white adults, respectively, on average $700 and $500 higher than in the standard model. ICERs for adding the new treatment were consistently lower for non-Hispanic Black adults compared to non-Hispanic white adults across both models.

### 3.4 Sensitivity analysis

#### 3.4.1 Alternative disease characteristics

Under modified disease characteristics, the cumulative incidence of sickness was 4% (95% CI: 3%, 4%) and 4% (95% CI: 4%, 4%) for non-Hispanic Black and non-Hispanic white adults, respectively (Table S6). As a result of fewer simulated individuals being affected by the sickness, adding the new treatment to standard of care did not result in improvements in life expectancy for non-Hispanic Black adults in either the standard model or the social factors framework.

However, consistent with our main analysis, under the social factors framework, adding the new treatment widened disparities in the number of years spent sick by 0.4 years and the fraction of sick years spent on treatment by 5 percentage points between non-Hispanic Black and non-Hispanic white adults. This result was not observed in the standard model.

Finally, ICERs for adding the new treatment were less favorable when using the social factors framework compared to the standard model for both groups. As in the main analysis, increases in ICERs were more pronounced for non-Hispanic Black adults than for non-Hispanic white adults under these alternative disease characteristics (Table S7, Table S8).

### 3.4.2 Differential healthcare costs by insurance status

When considering higher healthcare costs for individuals without insurance, the incremental healthcare costs of adding the new treatment increased for both groups (Table S9, Table S10) under the social factors framework. These increases in costs were larger for non-Hispanic Black adults, reflecting their lower rates of health insurance coverage compared with non-Hispanic white adults.

## 4 Discussion

Our findings underscore the importance of incorporating social factors in health policy simulation models. In the simplified decision problem, we found that failing to consider social factors produced biased estimates of benefits, healthcare utilization, costs, and health disparities. Results from our social factors framework suggest that non-Hispanic white adults experienced greater gains in health outcomes from the new treatment, which exacerbated health disparities between non-Hispanic Black and non-Hispanic white adults. Had our framework not been implemented, we may have incorrectly concluded that this treatment would have no impact on health disparities between the two racial/ethnic groups.

These differences can be attributed to the inclusion of a key social factor, health insurance coverage. In the social factors framework, we assumed that health insurance status impacted both baseline mortality rates and access to regular healthcare. This first assumption reflected evidence from prior studies that uninsured individuals may face higher baseline mortality risks due to increased prevalence of underlying health conditions, inadequate access to healthcare for other conditions, and increased stress.^9,14^ The second assumption asserted that insurance status influenced disease-specific outcomes by affecting healthcare and treatment access.^11,12^ Due to racial/ethnic differences in health insurance coverage, with the social factors framework, non-Hispanic Black adults spent a smaller proportion of their sick years receiving treatment compared to non-Hispanic white adults. Reduced treatment access diminished the potential benefits of implementing the new treatment for the non-Hispanic Black group. Our social factors framework highlighted inequities in treatment access and the downstream effect on health outcomes, which were overlooked in the standard model.

The cost-effectiveness analysis further emphasized these results. Adding the new treatment to standard of care yielded smaller QALY gains in the social factors framework compared to the standard model for both racial/ethnic groups, with reductions being more pronounced for non-Hispanic Black adults. Despite these diminished QALY gains, ICERs remained largely consistent between the two models. Reduced access to treatment resulted in lower incremental QALYs and costs, while keeping ICERs relatively consistent. It is possible that ICERs may diverge when incorporating the social factors framework in other scenarios involving high up-front costs for diagnosis and treatment initiation, attenuated treatment effectiveness over time, or changes in quality-of-life weights that vary across different health system states. Results from the simplified decision problem highlight two limitations of traditional cost-effectiveness analyses in addressing health disparities. First, as demonstrated by the standard model, these methods often assume homogeneous and guaranteed access to healthcare within the target population, thereby ignoring individuals in the population who lack access to healthcare. When we accounted for real-world heterogeneity in healthcare access using the social factors framework, the projected incremental costs and QALYs changed. Second, conventional cost-effectiveness metrics do not capture distributional effects on costs and health outcomes, as ICERs cannot reflect for whom the benefits were the largest. These findings reinforce that traditional cost-effectiveness analyses alone may be insufficient to inform decision-making on interventions with health equity implications.

Health policy simulation models have faced criticism for their lack of effectiveness in addressing health disparities.^15^ The Second Panel on Cost-effectiveness and Medicine discussed the importance of assessing who within a population benefits from health interventions, often referred to as the distributional impact.^16^ To fill this gap, several approaches have been developed, including equity impact analyses, extended cost-effectiveness analyses, and distributional cost-effectiveness analyses.^17–20^ However, these methods do not provide recommendations for how to explicitly model social factors and their relationship to disease progression or health system utilization. In particular, distributional cost-effectiveness analyses offer a framework for navigating equity-efficiency trade-offs in decision-making. While the social factors framework does not address these trade-offs, it can improve estimates of the distribution of costs and benefits accrued to individuals across dimensions of disparities required in extended and distributional cost-effectiveness analyses. Additionally, other research has considered including social determinants of health in health policy modeling,^21^ but to our knowledge, no formal framework exists for integrating these factors into health policy simulation models.

Our social factors framework has several limitations. First, the limited availability and quality of individual-level data on social drivers of health can pose a nontrivial challenge.^22^ These data are often incomplete, requiring researchers to rely on location-based proxies, such as administrative or geographic units (e.g., census tract, postal codes, regional divisions).^23^ Building models to address health equity concerns will benefit from greater investment in high-quality, individual-level data on social factors.^24,25^ Second, incorporating social factors into health policy simulation models requires careful consideration of their complex causal pathways.^26–28^ This includes longitudinal, time-dependent processes between social factors, healthcare utilization, and health outcomes. In our simplified decision problem, we assumed that insurance access has an effect on utilization and outcomes but did not incorporate, for example, the later effects of health outcomes on insurance status. Third, it may be necessary to consider heterogeneity in treatment effectiveness or disease-specific mortality risks, which may vary directly or indirectly due to social drivers of health, family history, or genetic factors. Finally, it may be challenging to apply in certain types of health policy simulation models. For example, state-transition cohort models are generally used for simulating homogeneous cohorts. Therefore, incorporating our framework to account for heterogeneous social factors would require stratification.

In our simplified decision problem, we demonstrated that ignoring heterogeneous health insurance coverage led to the incorrect conclusion that the new treatment did not exacerbate health disparities, potentially resulting in misinformed policy recommendations. By incorporating social factors, our framework enables the evaluation of an intervention’s social impact before implementation and facilitates comparisons between clinical and social interventions. We recommend our approach as a valuable tool for health policy decision-analytic models, particularly when evaluating health interventions with equity considerations.

## Acknowledgments

We thank Malcolm Barrett for his valuable contributions in code review and the Health Policy Data Science Lab for feedback on figure design.

## Ethical considerations

This study did not require Institutional Review Board approval because all data were publicly available and did not involve human participants.

## Consent to participate

Not applicable

## Patient consent

Not applicable

## Consent to publish

Not applicable

## Data availability

The Python code to reproduce our results is available at: https://github.com/StanfordHPDS/social_factors_microsim.

## Supplement Tables

**Table S1:** Modified parameters for sensitivity analysis alternative disease characteristics ^4^.

| Parameter |  | Base Case | Sensitivity Analysis |
| --- | --- | --- | --- |
| *Disease natural history annual transition probabilities* |  |  |  |
| Healthy | Sick | 0.05 | 0.001 |
| *Increased mortality risk* |  |  |  |
| Hazard ratio of death<br>(sick vs. healthy) |  | 4.0 | 6.0 |
| *Cost-effectiveness analysis parameters* |  |  |  |
| State-specific<br>quality-of-life weights | Sick | 0.7 | 0.5 |
| Annual state-specific<br>costs | Sick | \$500 | \$5,000 |

**Table S2:** Modified parameters for sensitivity analysis of differential healthcare costs by insurance status ^5^.

| Parameter |  | Base Case | Sensitivity Analysis |
| --- | --- | --- | --- |
| *Cost-effectiveness analysis parameters* |  |  |  |
| Annual state-specific<br>costs | Sick (insured) | \$500 | \$500 |
| | Sick (uninsured) | \$500 | \$2,000 |
| Annual treatment<br>costs | New treatment<br>(insured) | \$6,000 | \$6,000 |
| Annual treatment<br>costs | New treatment<br>(uninsured) | \$6,000 | \$24,000 |

**Figure S1:**
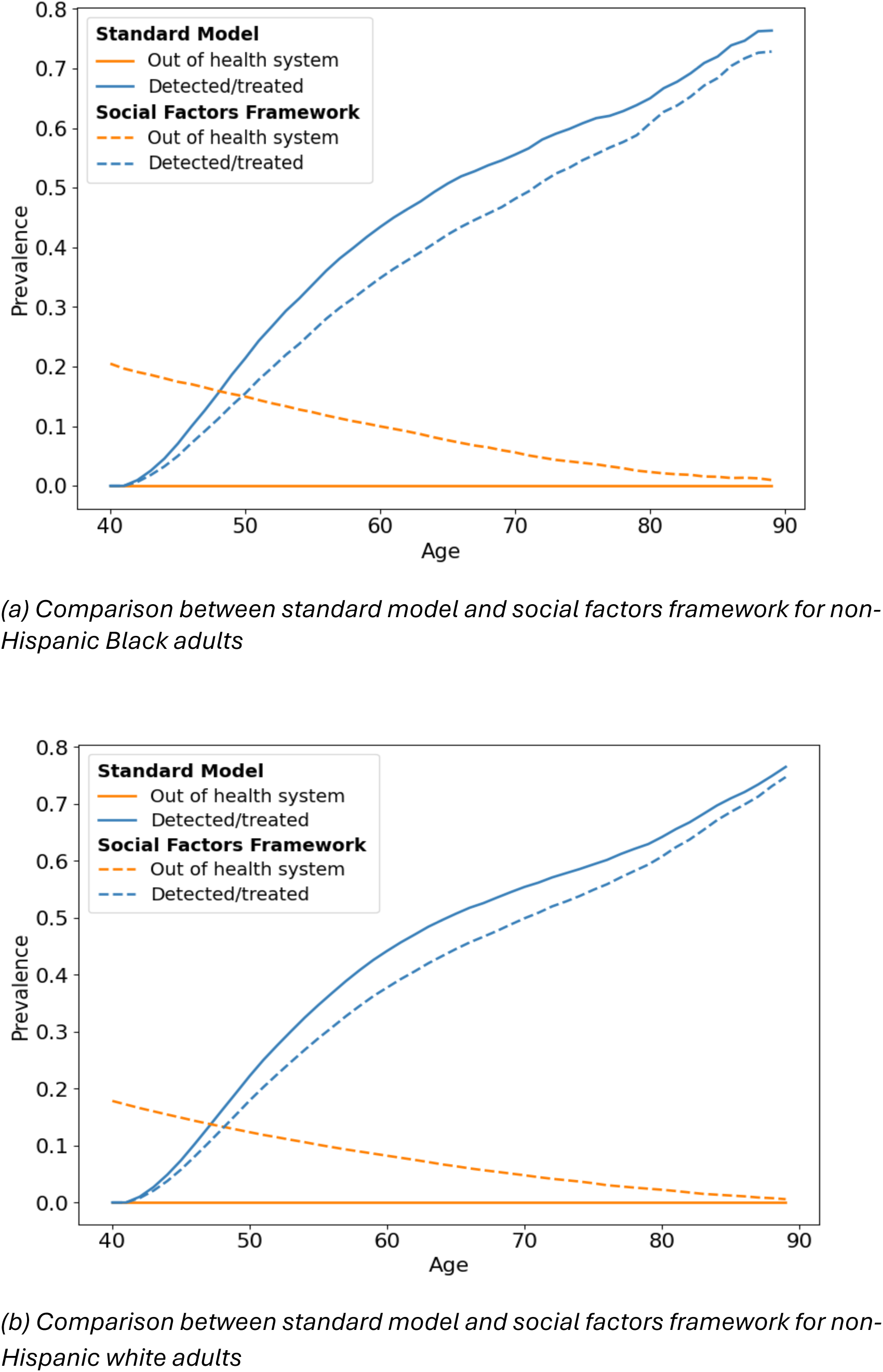
Model trace of simulated cohort across health system utilization states over 60 years.

**Table S3:** Cost and QALY outcomes for non-Hispanic Black and non-Hispanic white adults.

| Parameter | Group | Standard Model |  | Social Factors Framework |  |
| --- | --- | --- | --- | --- | --- |
|  |  | *Standard of Care* | *New Treatment* | *Standard of Care* | *New Treatment* |
| QALYs | Non-Hispanic Black | 16.5 [<0.1] | 17.2 [<0.1] | 16.2 [<0.1] | 16.7 [<0.1] |
|  | Non-Hispanic white | 17.8 [<0.1] | 18.5 [<0.1] | 17.6 [<0.1] | 18.1 [<0.1] |
|  | Difference between groups | 1.3 [<0.1] | 1.2 [<0.1] | 1.4 [<0.1] | 1.4 [<0.1] |
| Costs | Non-Hispanic Black | \$5,800 [<\$100] | \$37,200 [\$300] | \$5,400 [<\$100] | \$29,000 [\$300] |
| | Non-Hispanic white | \$6,800 [<\$100] | \$43,200 [\$100] | \$6,400 [<\$100] | \$36,700 [\$100] |
| | Difference between groups | \$1,000 [<\$100] | \$6,000 [\$300] | \$1,100 [<\$100] | \$7,700 [\$300] |

**Table S4:**
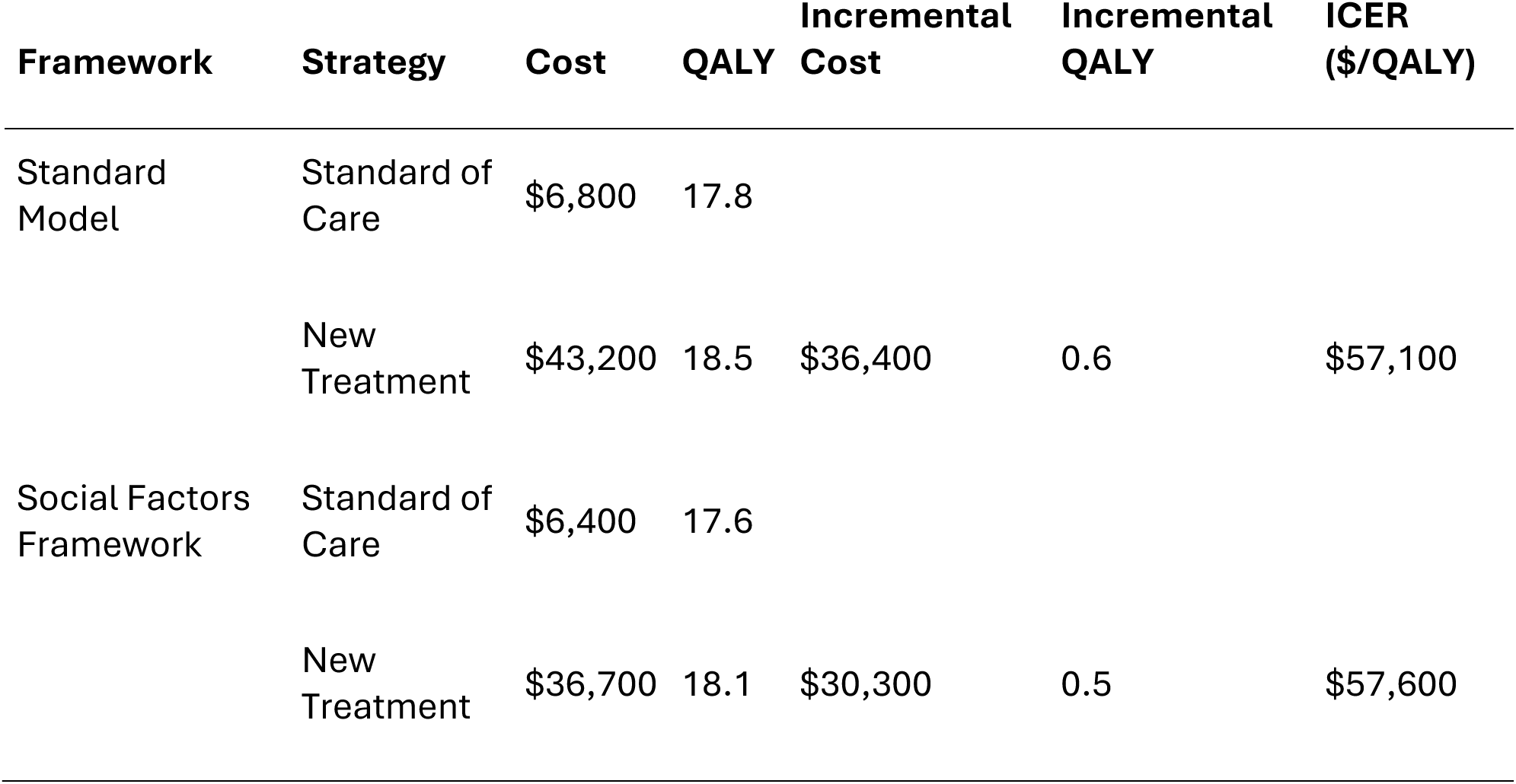
Cost-effectiveness results for non-Hispanic white adults.

| Framework | Strategy | Cost | QALY | Incremental Cost | Incremental QALY | ICER (\$/QALY) |
| --- | --- | --- | --- | --- | --- | --- |
| Standard Model | Standard of Care | \$6,800 | 17.8 | | | |
| | New Treatment | \$43,200 | 18.5 | \$36,400 | 0.6 | \$57,100 |
| Social Factors Framework | Standard of Care | \$6,400 | 17.6 | | | |
| | New Treatment | \$36,700 | 18.1 | \$30,300 | 0.5 | \$57,600 |

**Table S5:** Cost-effectiveness results for non-Hispanic Black adults.

| Framework | Strategy | Cost | QALY | Incremental Cost | Incremental QALY | ICER (\$/QALY) |
| --- | --- | --- | --- | --- | --- | --- |
| Standard Model | Standard of Care | \$5,800 | 16.5 | | | |
| | New Treatment | \$37,200 | 17.2 | \$31,400 | 0.7 | \$46,200 |
| Social Factors Framework | Standard of Care | \$5,400 | 16.2 | | | |
| | New Treatment | \$29,000 | 16.7 | \$23,600 | 0.5 | \$46,900 |

**Table S6:**
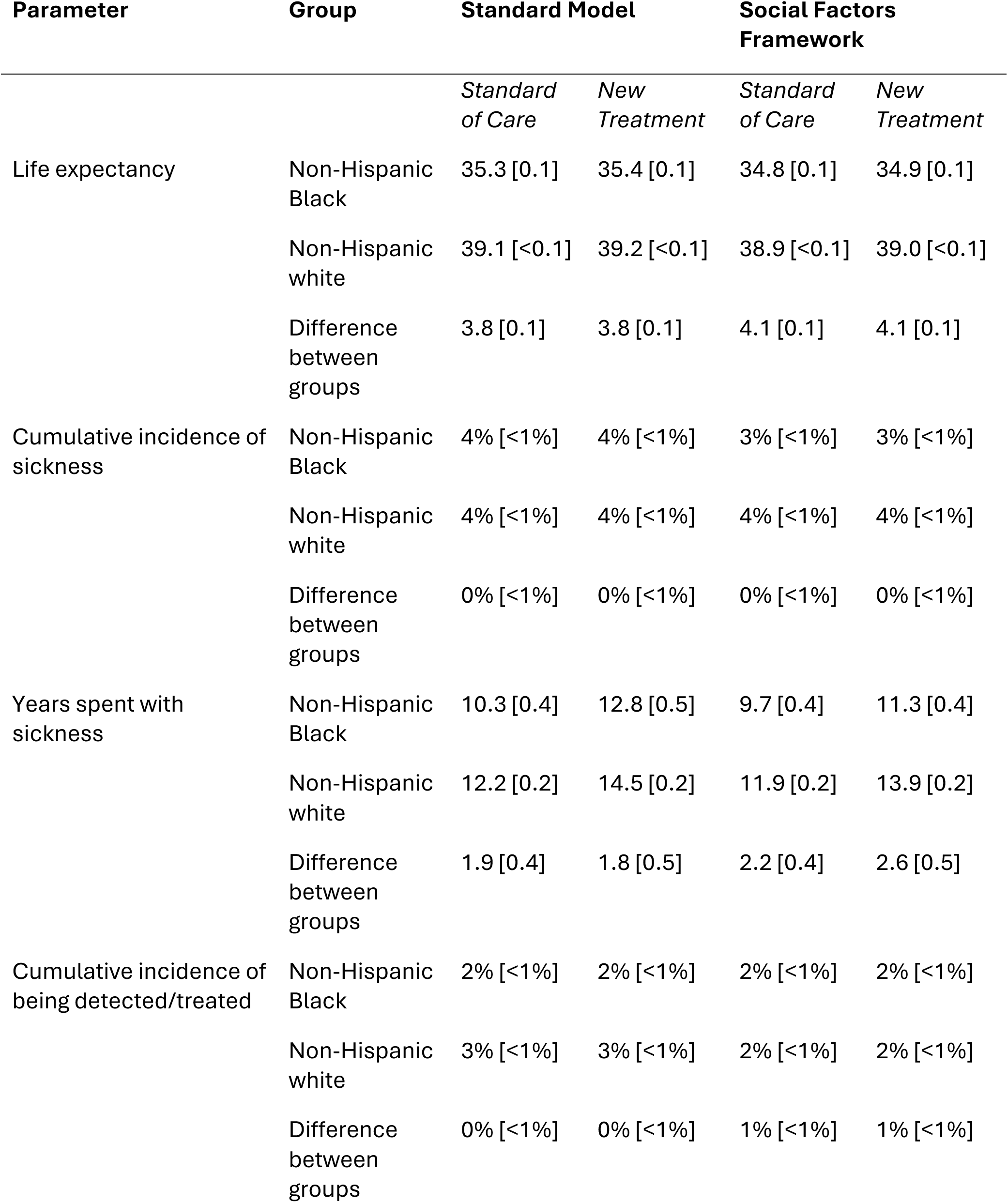
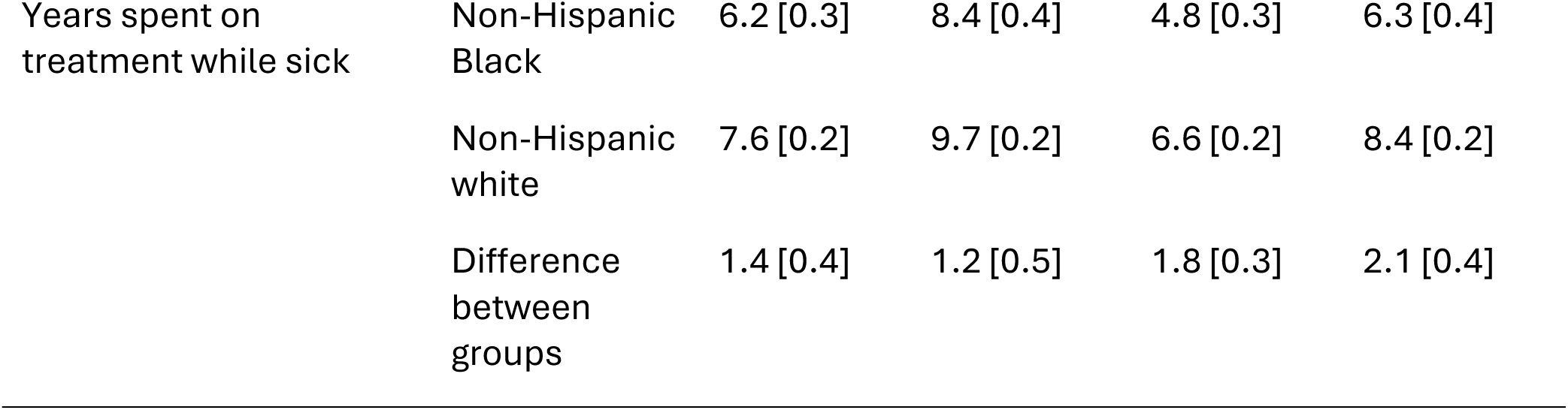
Main outcomes in sensitivity analysis of alternative disease characteristics.

**Table S7:** Cost-effectiveness results for non-Hispanic white adults (alternative disease characteristics)

| Framework | Strategy | Cost | QALY | Incremental Cost | Incremental QALY | ICER (\$/QALY) |
| --- | --- | --- | --- | --- | --- | --- |
| Standard Model | Standard of Care | \$3,500 | 22.5 | | | |
| | New Treatment | \$4,700 | 22.5 | \$1,200 | 0.0 | \$76,400 |
| Social Factors Framework | Standard of Care | \$3,400 | 22.4 | | | |
| | New Treatment | \$4,400 | 22.4 | \$1,000 | 0.0 | \$75,500 |

**Table S8:** Cost-effectiveness results for non-Hispanic Black adults (alternative disease characteristics)

| Framework | Strategy | Cost | QALY | Incremental Cost | Incremental QALY | ICER (\$/QALY) |
| --- | --- | --- | --- | --- | --- | --- |
| Standard Model | Standard of Care | \$3,100 | 21.0 | | | |
| | New Treatment | \$4,100 | 21.0 | \$1,000 | 0.0 | \$61,400 |
| Social Factors Framework | Standard of Care | \$3,000 | 20.8 | | | |
| | New Treatment | \$3,700 | 20.8 | \$700 | 0.0 | \$70,100 |

**Table S9:** Cost-effectiveness results for non-Hispanic white adults (differential costs by insurance status)

| Framework | Strategy | Cost | QALY | Incremental Cost | Incremental QALY | ICER (\$/QALY) |
| --- | --- | --- | --- | --- | --- | --- |
| Standard Model | Standard of Care | \$6,800 | 17.8 | | | |
| | New Treatment | \$43,200 | 18.5 | \$36,400 | 0.6 | \$57,100 |
| Social Factors Framework | Standard of Care | \$7,800 | 17.6 | | | |
| | New Treatment | \$40,200 | 18.1 | \$32,400 | 0.5 | \$61,700 |

**Table S10:** Cost-effectiveness results for non-Hispanic Black adults (differential costs by insurance status)

| Framework | Strategy | Cost | QALY | Incremental Cost | Incremental QALY | ICER (\$/QALY) |
| --- | --- | --- | --- | --- | --- | --- |
| Standard Model | Standard of Care | \$5,800 | 16.5 | | | |
| | New Treatment | \$37,200 | 17.2 | \$31,400 | 0.7 | \$46,200 |
| Social Factors Framework | Standard of Care | \$7,300 | 16.2 | | | |
| | New Treatment | \$33,900 | 16.7 | \$26,600 | 0.5 | \$52,700 |

## Footnotes

1 Individuals in the health system are undetected and untreated for the sickness.

2 In the standard model, all individuals are initiated within the health system.

3 Parameters without citations are assumed parameters.

4 We estimated the proportion of our cohort in the health system state using the National Health and Nutrition Examination Survey question for routine place to go for healthcare.

4 Parameters were modified for both standard model and model with social factors framework

5 Parameters were only modified for the model with the social factors framework

